# Acceptability of cannabidiol as a treatment for people at clinical high risk for psychosis

**DOI:** 10.64898/2026.03.05.26347694

**Authors:** Dominic Oliver, Edward Chesney, Phoebe Wallman, Andrés Estradé, Matilda Azis, Umberto Provenzani, Stefano Damiani, Antonio Melillo, Olivia Hunt, Shubhi Agarwala, Amedeo Minichino, Peter J. Uhlhaas, Philip McGuire, Paolo Fusar-Poli

**Author notes:** **Corresponding author:** Dr Dominic Oliver, POWIC Building, Department of Psychiatry, University of Oxford, Warneford Lane, Oxford OX3 7JX, UK. These authors contributed equally (Philip McGuire, Paolo Fusar-Poli).

## Abstract

**Background:** At present, there are no approved pharmacological treatments for people at clinical high risk for psychosis (CHR-P). We sought to assess the acceptability of cannabidiol (CBD): a promising candidate treatment for this population.

**Methods:** CHR-P individuals completed a survey which assessed their views on the acceptability of CBD, its expected effectiveness and side effects, and on formulation preferences.

**Results:** The sample comprised 55 CHR-P individuals (24.3 years and 69% female). Most (91%) were familiar with CBD, and had previously used cannabis (64%), and around half (42%) had tried over-the-counter CBD. 75% were willing to take CBD as an intervention for mental health problems. Most participants anticipated fewer side effects with CBD than with existing medications, and preferred tablet or capsule formulations over liquids.

**Discussion:** CBD is perceived as a highly acceptable treatment among CHR-P individuals.

## INTRODUCTION

The onset of psychosis is usually preceded by a detectable clinical high risk for psychosis (CHR-P) state (1,2), which is characterised by attenuated psychotic symptoms. Around 20% of CHR-P individuals will develop psychosis within two years (3,4).

Clinical intervention during the CHR-P phase may reduce the severity of presenting symptoms, and has the potential to delay or prevent the onset of psychosis (5). However, meta-analyses indicate that there is no robust evidence that any pharmacological or psychological intervention is superior to needs-based interventions in reducing attenuated positive symptoms or the risk of transition to psychosis (6). At present, there are no licensed pharmacological interventions for CHR-P individuals (7).

Data from clinical trials suggest that cannabidiol (CBD) can reduce severity of psychotic symptoms in patients with psychosis (8,9), and in people at CHR-P (10).

A meta-analysis of randomised, placebo-controlled trials found that CBD has a few adverse effects and high tolerability (11). This relatively benign side effect profile contrasts with that of antipsychotic medications (12), which are often associated with weight gain, Parkinsonian symptoms, sexual dysfunction and sedation. In addition, CBD appears to be perceived as less stigmatising than antipsychotic mediations, partly because it is a constituent of cannabis, and because it is available as an over-the-counter health supplement (13,14). In a recent survey of people with psychotic disorders, CBD had a very high acceptability, with 86% saying that they would be willing to take it as a treatment (15). Its acceptability as an intervention in CHR-P individuals has yet to be formally assessed.

The aim of this study was therefore to assess the acceptability of CBD as a potential intervention among people with CHR-P. We hypothesised that CBD would be perceived as a highly acceptable intervention, particularly in comparison to antipsychotic medications.

## METHODS

### Design and setting

This study was part of a larger research program testing a digital screening method for emerging disorders in the community: E-Detection Tool for Emerging Mental Disorders (ENTER; IRAS: 298213, Istituti di Ricovero e Cura a Carattere Scientifico San Matteo 0044135/22). ENTER’s details are presented in a separate publication (16). Assessments were performed at King’s College London, the University of Glasgow (both UK) and University of Pavia (Italy).

### Participants

Individuals aged 12 to 35 were recruited through a comprehensive outreach campaign, which included flyers, digital advertisements on social media (Instagram, Facebook and TikTok) and university newsletters. Individuals were invited to access the ENTER website and to consent to the subsequent study procedures. The included sample comprised participants who scored ≥6 on the online Prodromal Questionnaire 16-item version (PQ-16) (17), and subsequently attended an in person interview where they were assessed with the Comprehensive Assessment for At-Risk Mental States (CAARMS) (18) and completed the CBD acceptability questions. A subset of these participants (CHR-B) were additionally assessed with the Structured Interview for Bipolar At Risk States (SIBARS) (19,20). All participants gave written informed consent to participate and received financial compensation for participating in the study. Data were collected between January 2023 and December 2025.

### CBD acceptability

Participants answered an 15-item questionnaire via interview (see Supplement). We provided a brief description of CBD before sections exploring (i) prior experience of cannabis and CBD, (ii) willingness to try interventions for mental health, (iii) expected treatment effects of CBD, (iv) expected side effects and (v) preferences for formulations.

### Statistical analysis

Analyses for the primary analyses were restricted to participants who met CHR-P criteria. Willingness to trial CBD was analysed according to demographic and clinical variables, tested with Fisher’s exact test. Fisher’s exact test was used as the expected values in contingency tables were small. When there were more than two categories, a pairwise Fisher’s exact test was used to examine all possible contrasts. Intervention preferences were compared using pairwise Wilcoxon signed rank test with Holm continuity correction.

As secondary analyses, we assessed willingness to consider any intervention for their mental health and CBD in participants who met CHR-B criteria.

The threshold for statistical significance was set as p<0.05. All analyses were completed using R version 4.4.2.

## RESULTS

The demographic and clinical characteristics of the included participants (n=55) are shown in Table 1. The mean age was 24.2 (SD=3.7) years and 66% of the participants were female. 91% had already heard of CBD, 48% had used an over-the-counter CBD product. Most participants (64%) had previously used cannabis.

**Table 1.**
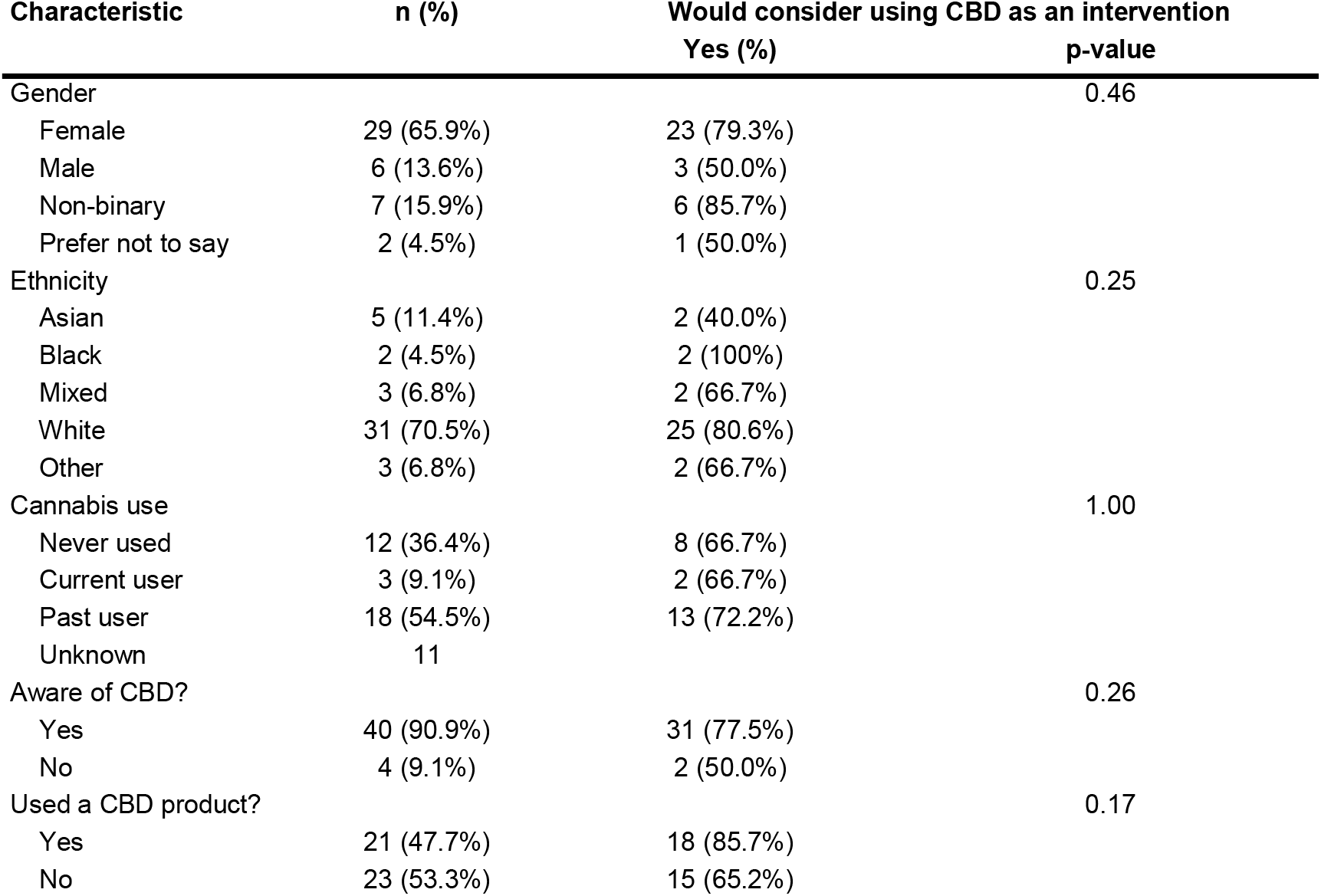
Proportion of participants who would consider taking CBD as an intervention for their mental health according to demographic and clinical characteristics.

| Characteristic | n (%) | Would consider using CBD as an intervention<br>Yes (%) | p-value |
| --- | --- | --- | --- |
| Gender |  |  | 0.46 |
| Female | 29 (65.9%) | 23 (79.3%) |  |
| Male | 6 (13.6%) | 3 (50.0%) |  |
| Non-binary | 7 (15.9%) | 6 (85.7%) |  |
| Prefer not to say | 2 (4.5%) | 1 (50.0%) |  |
| Ethnicity |  |  | 0.25 |
| Asian | 5 (11.4%) | 2 (40.0%) |  |
| Black | 2 (4.5%) | 2 (100%) |  |
| Mixed | 3 (6.8%) | 2 (66.7%) |  |
| White | 31 (70.5%) | 25 (80.6%) |  |
| Other | 3 (6.8%) | 2 (66.7%) |  |
| Cannabis use |  |  | 1.00 |
| Never used | 12 (36.4%) | 8 (66.7%) |  |
| Current user | 3 (9.1%) | 2 (66.7%) |  |
| Past user | 18 (54.5%) | 13 (72.2%) |  |
| Unknown | 11 |  |  |
| Aware of CBD? |  |  | 0.26 |
| Yes | 40 (90.9%) | 31 (77.5%) |  |
| No | 4 (9.1%) | 2 (50.0%) |  |
| Used a CBD product? |  |  | 0.17 |
| Yes | 21 (47.7%) | 18 (85.7%) |  |
| No | 23 (53.3%) | 15 (65.2%) |  |

87% of participants were willing to receive treatment for their mental health problems, and 76% were willing to consider taking CBD (Table 1). Although 44% had concerns about taking a cannabis-based medicine, 72% of those who were current or past users of cannabis and 67% of never cannabis users were willing to try treatment with CBD (OR=1.2, 95%CI=0.20-7.2, p=1.0). Similarly, 87% of participants who had previously used over-the-counter CBD products were willing to try intervention with CBD, compared with 69% of those who had never used them (OR=3.0, 95%CI=0.64-19.2, p=0.20). There were no effects of gender or ethnicity on the likelihood of considering accepting CBD treatment (p>0.05).

A minority of the participants (n=15) indicated that they would not wish to take CBD as an intervention. Some did not want to take any kind of medication (n=6). Others declined because CBD is derived from cannabis (n=4), because they felt there was a lack of evidence for its efficacy (n=3), due to concerns about mental health-related side effects (n=1), or because it was not ‘natural’ (n=1). Of those willing to take CBD, most (62%) were willing to take it for two years or more. 7% were willing to take it for one year, 12% for six months, and 12% for three months or less. 74% thought that their symptoms would improve with CBD, while only 2% thought it would make their symptoms worse.

### Intervention comparison

28 participants ranked their preferences for different classes of treatment: anti-anxiety medications, antidepressants, antipsychotics, CBD and talking therapies. CBD had a median rank of 3, significantly higher than the least popular (antipsychotics; median rank=5, p<0.001). There were no significant differences between the rankings for CBD and talking therapies, anti-anxiety medications and antidepressants (p>0.21).

Most participants wanted CBD to help with their anxiety (77%), followed by sleep problems (50%), mood dysfunction (40%), and unusual experiences (23%) (Figure 1).

**Figure 1.**
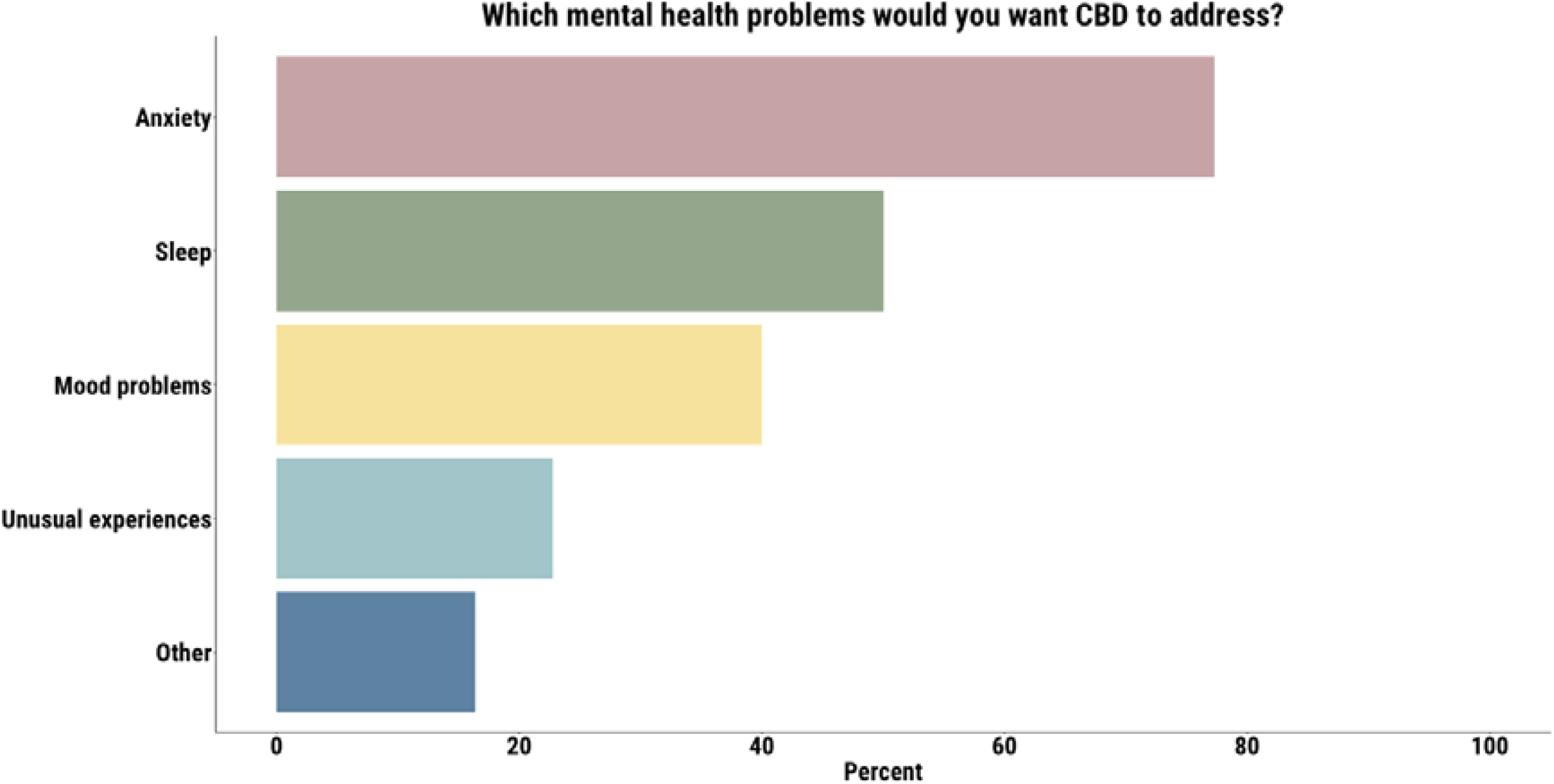
Anxiety was the most common symptom that participants wanted CBD to address

### Side effects

44 participants answered questions about side effects. 44% said that they thought that CBD would not have any side effects. When asked to compare CBD with their current medication (antidepressants and anti-anxiety medication), 73% of participants thought that CBD would have more tolerable side effects, whereas 8% thought that it would have worse side effects (Figure 2). When asked to compare CBD with talking therapies, 35% thought that CBD would have more tolerable side effects, and 24% thought that it would have worse side effects (Figure 2).

**Figure 2.**
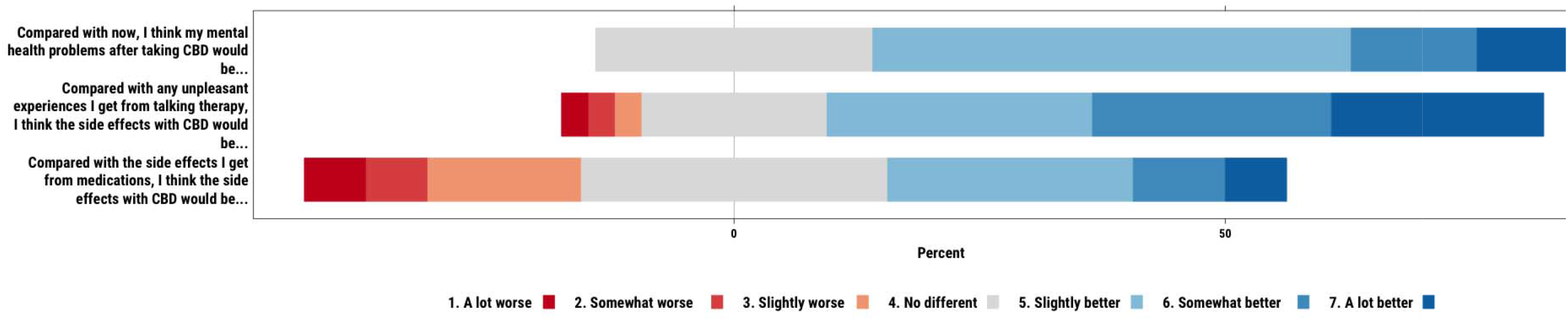
Participants’ expectations regarding the efficacy and side effects of CBD

The anticipated side effects for CBD included drowsiness (11%), stomach upset (9%), paranoia (5%), anxiety (2%), intoxication (2%), hearing or seeing things that aren’t really there (2%), feeling sick (2%), feeling tired (2%) and unusual beliefs (2%). No participant endorsed blurred vision, constipation, dizziness, dry mouth, feeling hungry, loss of sex drive, muscle twitches and spasms, pain relief, feeling relaxed/calm or shakiness/trembling as expected side effects.

### Formulation

Those open to using CBD were then asked about the acceptability of different formulations. Of the 34 participants who provided a response, 77% indicated that they would be willing to take a capsule, 73% a tablet, and 61% a liquid. When asked for their preferred formulation, 35% chose a tablet, 28% a capsule, 13% a liquid, and 9% chose edibles (Figure 3).

**Figure 3.**
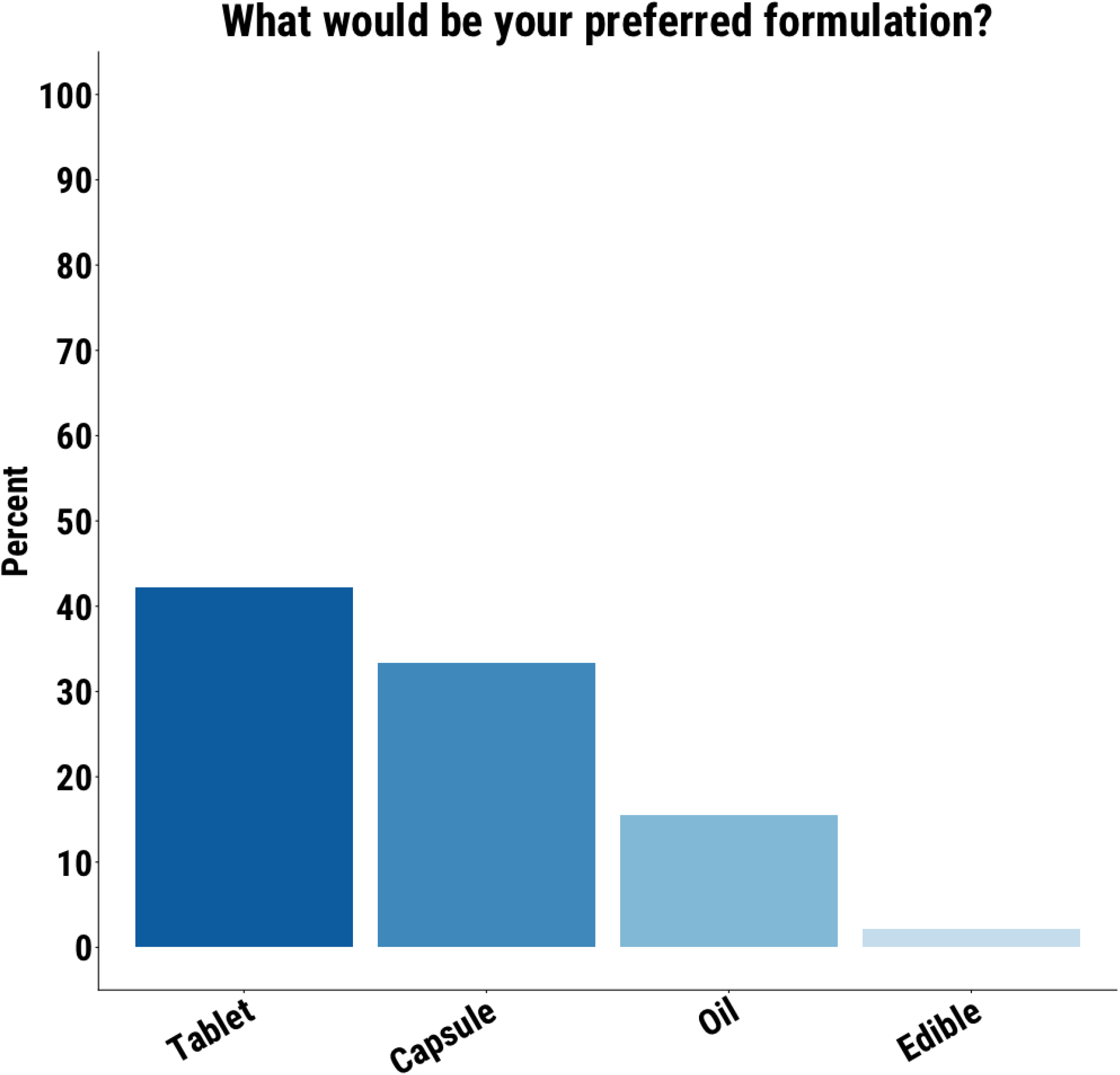
Tablet was the preferred formulation for the largest proportion of participants

### CHR-B

36 participants met CHR-B criteria. The mean age was 24.4 (SD=4.7) years and 72% of the participants were female, 19% were male and 8% were non-binary. 86% had already heard of CBD, 44% had used an over-the-counter CBD product. Most participants (63%) had previously used cannabis.

86% of participants meeting CHR-B criteria were willing to take an intervention for their mental health problems. 75% were willing to consider using CBD to treat their mental health problems.

## DISCUSSION

This study examined the acceptability of CBD as an intervention among people at clinical high risk for psychosis. The main finding was that most viewed CBD as a highly acceptable intervention, with 75% of participants willing to take it for their mental health problems. Most participants also had positive expectations about the benefits and side-effects of CBD relative to existing interventions.

The high proportion of CHR-P individuals willing to take CBD is consistent with previous findings in patients with psychosis (15). Moreover, most of the CHR-P participants who were willing to be treated with CBD were prepared to take it for two years or more. Because the maximal period of risk for psychosis in the CHR-P population is over the first two years (21–23), ideally, treatments intended to prevent the later onset of psychosis would be delivered over this period. For example, in an ongoing trial of CBD as a preventive intervention for psychosis, the duration of treatment is two years (24). Although all previous trials of CBD in psychosis have been short-term, the findings from the present study suggest that long term treatment in CHR-P individuals is feasible.

In a previous study which investigated CBD’s acceptability in patients with psychosis, prior use of cannabis and over-the-counter CBD products were associated with greater willingness to consider CBD as an intervention (15). However, we did not observe these associations in CHR-P participants. This may in part be due to low statistical power within our sample. Despite half of participants having some reservations about a cannabis-based medicine, most were still willing to take CBD. This suggests that these concerns only put a small minority off accepting CBD, with only four participants citing this as the specific reason for declining. Providing psychoeducation about CBD’s effects and safety profile to people who are being offered it as an intervention may be useful to reduce any minor concerns.

Participants generally expected that CBD would improve mental health symptoms and thought that its side-effect profile was better than that of their current medications (antidepressants and anti-anxiety medication). While these expectations may support willingness to take CBD, they also highlight the importance of managing treatment expectations as they can modulate treatment response on psychiatric outcomes (25,26) and may contribute to the placebo effect, which can be significant in CHR-P individuals (27,28). It is thus possible that the relatively positive expectations about treatment with CBD could enhance its effectiveness in real-world practice. If an individual believes that CBD is an effective treatment, it may reduce anxiety and alleviate stress, which in turn may reduce the risk of adverse long-term outcomes as they may contribute to psychosis risk (29,30).

When directly compared with other intervention options, CBD was ranked more favourably than antipsychotic medications. It did not differ significantly in acceptability from talking therapies, antidepressants or anti-anxiety medications, suggesting that it is perceived as comparable to commonly prescribed first-line psychological and pharmacological interventions in early detection settings. This is encouraging, as a small clinical trial suggests that CBD can reduce symptom severity in people with CHR-P (10). Participants expected that CBD would mostly have fewer unpleasant effects than talking therapies and better side effect profiles compared to their current medications. Participants’ preferences for CBD to primarily help with anxiety, sleep and mood (rather than attenuated psychotic symptoms), as well as similar acceptability in participants meeting CHR-B criteria, further support its perceived role as a transdiagnostic intervention targeting common and distressing symptoms in early stages of the disorder. It is important, therefore, that trials of CBD in CHR-P populations also assess these outcomes. These findings, alongside the possible anxiolytic effects of CBD (31), underscore the importance of framing CBD not solely as a potential intervention to prevent the onset of psychosis, but also to treat current symptoms.

Preferences for tablets and capsules indicate that conventional pharmaceutical formulations may be more acceptable over oils. However, the only currently approved formulation of CBD is an oil-based formulation (32,33). Ongoing work to develop alternative CBD formulations is therefore warranted (34).

Several limitations should be acknowledged. We measured prospective acceptability, which may mean that many CHR-P individuals are willing to try CBD, it does not mean that they will adhere to treatment over the long-term (35). The cross-sectional design and modest sample size limited subgroup analyses and potential drivers of CBD acceptability. We only included participants from sites in the UK and Italy so these findings may not generalise to other international settings, particularly settings with different legislation regarding the legal status of cannabis.

In conclusion, CBD is perceived as a highly acceptable potential intervention among young people with CHR-P, emphasising the potential of CBD as an intervention in this population.

## Supporting information

CBD Questionnaire

## Data Availability

All data produced in the present study are available upon reasonable request to the authors.

## Acknowledgements

DO, AM and PM are supported by the National Institute for Health and Care Research (NIHR) Oxford Health Biomedical Research Centre. EC is funded by a Clinical Lectureship from NIHR. PFP is supported by the NIHR Maudsley Biomedical Research Centre. The views expressed are those of the author(s) and not necessarily those of the NHS, the NIHR or the Department of Health. AM is supported by a Wellcome Trust Early Career Award (304693/Z/23/Z). PM is funded by the Wellcome Trust (STEP). PFP is supported by #NEXTGENERATIONEU (NGEU), funded by the Ministry of University and Research (MUR), National Recovery and Resilience Plan (NRRP), project MNESYS (PE0000006) – A Multiscale integrated approach to the study of the nervous system in health and disease (DN. 1553 11.10.2022).

DO, EC, PM and PFP conceptualised the study. PW, AE, MA, UP, SD, AM, OH and SA collected the data. DO ran the statistical analyses. All authors drafted, edited and approved the final version of the manuscript.

