## Supplementary material for "Acceptability of cannabidiol as a treatment for people at clinical high risk for psychosis": CBD Questionnaire

**Introduction**

Cannabidiol is a new medication which may be an effective treatment for mental health problems. It is made naturally by the cannabis plant but it isn’t intoxicating and won’t get you ‘high’. Unlike cannabis, studies have found CBD may help with unusual experiences like paranoia or hearing voices. Many people use it as a health supplement for problems such as stress, anxiety and pain. It has very few side effects, though may cause stomach upset. We have a few questions about perceptions of this treatment from people who may benefit from it in the future.

**Background**

1. If a treatment was available to help with your mental health problems, would you want to take it? [Yes/No]
2. Have you ever heard of cannabidiol (CBD) before? [Yes/No]
3. Have you ever used CBD before [Yes/No]

**Treatment effects**

4. If CBD was available as a medication to treat your mental health problems, would you want to take it? [Yes/No] [if no, specify]
5. How long would you be open to taking CBD for if it was available as a medication to treat your mental health problems? [3 months or less/6 months/1 year/2 years or longer]
6. Compared with now, I think my mental health problems after taking CBD would be… [a lot worse/slightly worse/somewhat worse/no different/slightly better/somewhat better/a lot better]
7. Which mental health problems would you want CBD to address? [Mood problems/Anxiety/Unusual experiences (e.g. hearing voices, feeling paranoid)/Sleep/Other - specify]

**Adverse effects**

8. Do you think CBD has any side effects? [Yes/No]
9. How severe do you think the side effects would be? [1 (Mild) 2 3 4 (Moderate) 5 6 7 (Severe)]
10. What do you think the side effects would be? [high/ relaxed/ drowsiness/ shaking and trembling/ weight gain/ restlessness/ muscle twitches and spasms – where your muscles shorten tightly and painfully/ blurred vision/ dizziness/ constipation/ loss of sex drive (libido)/ dry mouth/ stomach upset/ other – specify]
11. Compared with the side effects I get from medications, I think the side effects with CBD would be... [a lot worse/slightly worse/somewhat worse/no different/slightly better/somewhat better/a lot better]
12. Compared with any unpleasant experiences I get from talking therapy, I think the side effects with CBD would be... [a lot worse/slightly worse/somewhat worse/no different/slightly better/somewhat better/a lot better]
13. Do you have any concerns about using a cannabis-based medicine? [Yes/No] [if yes, specify]

**Formulation**

14. Would you be happy to take CBD as a… [tablet/capsule/oil/other]
15. Which form of CBD would you prefer to take?
